# COVID-19 third wave experience in India, a survey of 5971 adults

**DOI:** 10.1101/2022.04.26.22274273

**Authors:** Rajeev Jayadevan, Ramesh Shenoy, TS Anithadevi

**Affiliations:** Sunrise Hospital; Lisie Hospital Cochin India; Little Flower Hospital Angamaly Kerala India

## Abstract

**Background:** The third wave of the pandemic in India lasted from January till March 2022, and breakthrough infections were common. Third dose of vaccine was rolled out to priority groups in the beginning of 2022. There is no published information available about the clinical outcomes in this context.

**Aims:**

1. To assess the community level experience of the pandemic, with focus on the third wave and vaccination in India.
2. To describe the experience of the boosted and non-boosted population during the 3^rd^ wave.
3. To study the public perception about the precautionary (3^rd^) dose in India.

**Results:** Among 5971 respondents, 98.6% were vaccinated, 40% of whom had also received the 3^rd^ dose. Age range: 24% were below 40, 50% were 40-59, 26% were >60 years.

45% were women, 53% were healthcare workers.

COVID-19 was reported by 3361 (56%) respondents. Among those who reported COVID-19, 2311 (70%) were infected during the third wave. Severe symptoms occurred in <1%, while moderate severity was reported by 42%. Repeated bouts of infection were common; 15% of those with a history of COVID-19 had been infected at least twice. 44% of the respondents (2610/5971) did not report a history of COVID-19.

The third dose was taken by 2383 individuals, of whom 30% reported COVID-19 during the 3^rd^ wave. The boosted group also had higher N95 use, and a greater proportion of healthcare workers. Among those who did not take a 3^rd^ dose, 45% reported COVID-19 in the 3^rd^ wave. COVID-19 incidence was lower at 27% among those in this group who had recently received their second dose. Longer gap after the second dose correlated with higher chance of infection during 3^rd^ wave. Giving a 3^rd^ dose before a 6-month gap since the second dose did not make a difference in infection rate.

Covaxin and Covishield recipients had the same incidence of COVID-19 during the third wave.

While 35% of the respondents believed it was helpful, 65% of the respondents were either uncertain or disapproving of the benefit of a 3^rd^ dose.

**Conclusions:**

1. 30% of respondents who received a 3^rd^ dose went on to get COVID-19 during the 3^rd^ wave.
2. Younger adults were more likely to be affected during 3^rd^ wave.
3. Although severe disease was rare, 42% reported having symptoms of moderate severity that could temporarily incapacitate people, affecting their routine and productivity.
4. The proportion of different grades of severity was similar among all vaccinated people, regardless of whether they received a 3^rd^ dose.
5. Reinfections occurred in 15%, and were not always milder.
6. Among those who did not receive a 3^rd^ dose, 45% reported COVID-19 in the 3^rd^ wave. However, this group had lower use of N95 masks (50%) than the 3^rd^ dose group (68%) which may have reduced the overall protection.
7. The longer the gap after the second dose, the greater was the chance of reporting COVID-19.
8. People who received their second dose recently had the same incidence of third wave COVID-19 as following a 3^rd^ dose.
9. The 3^rd^ dose, given too close to the second dose, made no difference in the infection rate.
10. Covaxin and Covishield recipients had the same rate of COVID-19 in the third wave.
11. Although the respondents were 98.6% vaccinated at baseline, there was considerable uncertainty (65%) amongst them about the benefit of a 3^rd^ dose.

## Background

The third wave of the pandemic arrived in India in late December 2021 and subsided by March 2022. Unlike during the delta wave in 2021, this time the virus was met by a largely vaccinated adult population: most adults had received either Covishield (adenovirus vector) or Covaxin (inactivated virus). In addition, many individuals had already acquired immunity through natural infection.

In India, 3^rd^ dose of the same vaccine (technically a homologous booster dose) was authorized for healthcare and frontline workers and for people over the age of 60, on January 10, 2022.

Both these vaccines are known to generate an immune response when used as a homologous booster dose (1,2). However, there is no published information available on the clinical outcomes of this intervention in the Indian context.

With more waves expected in the future, and wider availability of 3^rd^ doses now to all above the age of 18, it is important to collect pertinent information about the overall community experience during the third wave. The survey was therefore done as the Omicron-driven third wave was winding down in India, after sufficient time had elapsed for the 3^rd^ dose to take effect among recipients. At the time of completion of the study, 20 million precautionary (booster) doses had already been administered, while 96.4% adults had received at least one dose and 82% had received two doses (3).

For ease of discussion, the term 3^rd^ dose will henceforth be used instead of precautionary dose.

## Aim

1. To understand the 3^rd^ wave experience at the community level, with regard to infection rate, severity, vaccination status, gap since vaccine dose and 3^rd^ dose uptake.
2. To describe the experience of the boosted and non-boosted population during 3^rd^ wave.
3. To study the public perception about 3^rd^ dose vaccination.

## Methods

A cross-sectional online survey was performed from 15 February till 10 March 2022, which included questions pertaining to 3rd wave experience in India. The survey was sent for the attention of all adults based in India who may or may not have had a history of COVID-19. Questions were formatted in a clear and binary fashion and were validated before the wide launch through email and social media platforms.

For instance, to classify people according to disease severity, as a direct interview was not possible, we carefully constructed a simple-to-use multiple-choice question that had discriminatory power (3). Details are given in figure 12.

The questionnaire was kept as short and easy as possible to achieve the balance of generating pertinent unambiguous data, while enhancing survey referrals through a positive user experience. Incomplete responses could not be submitted. Only one response was possible from a single user.

The response was enthusiastic, with 5971 adults completing the survey. Provision was provided to add descriptive comments if the respondent felt the need to. This section also received considerable response, with as many as 795 people sending their comments.

The percentage of vaccinated respondents (98.6%) in the survey closely matched that of vaccinated adults in India at that time (96.4%). There was no clustering of unusual responses, indicating that the participation was legitimate and balanced, from individuals of varied demographic backgrounds across the country rather than focussed groups of people.

The data presented in the study were exclusively obtained through the online survey.

### Statistical Analysis

Descriptive statistics was used to present all outcomes. Binary and categorical variables were presented using counts and percentages. For the comparison of categorical variables, either chi square or Fisher’s exact test were used. All the data were entered in Microsoft excel and analyzed using SPSS version 20.00.

## Results

### 1. Profile of respondents

The survey was completed by 5971 adults based in India, 56% of whom reported a history of COVID-19. A substantial number (3180, 53%) worked in healthcare. Women comprised 45% of respondents. There was a wide range of age distribution; 50% belonged to the age group of 40-59, 24% were below 40, while 26% were 60 and above.

Among the 5971 survey respondents, the third dose was taken by 2383 individuals, of whom 1701 (71%) were healthcare workers.

### 2. Incidence of COVID-19 among respondents

39% (2311/5971) were infected during the third wave, while 18% (1050/5971) were infected during previous waves. Younger age groups were more likely to report COVID-19 during the 3^rd^ wave (see figure 2). 44% of the respondents (2610/5971) did not report a history of COVID-19.

**Figure 1:**
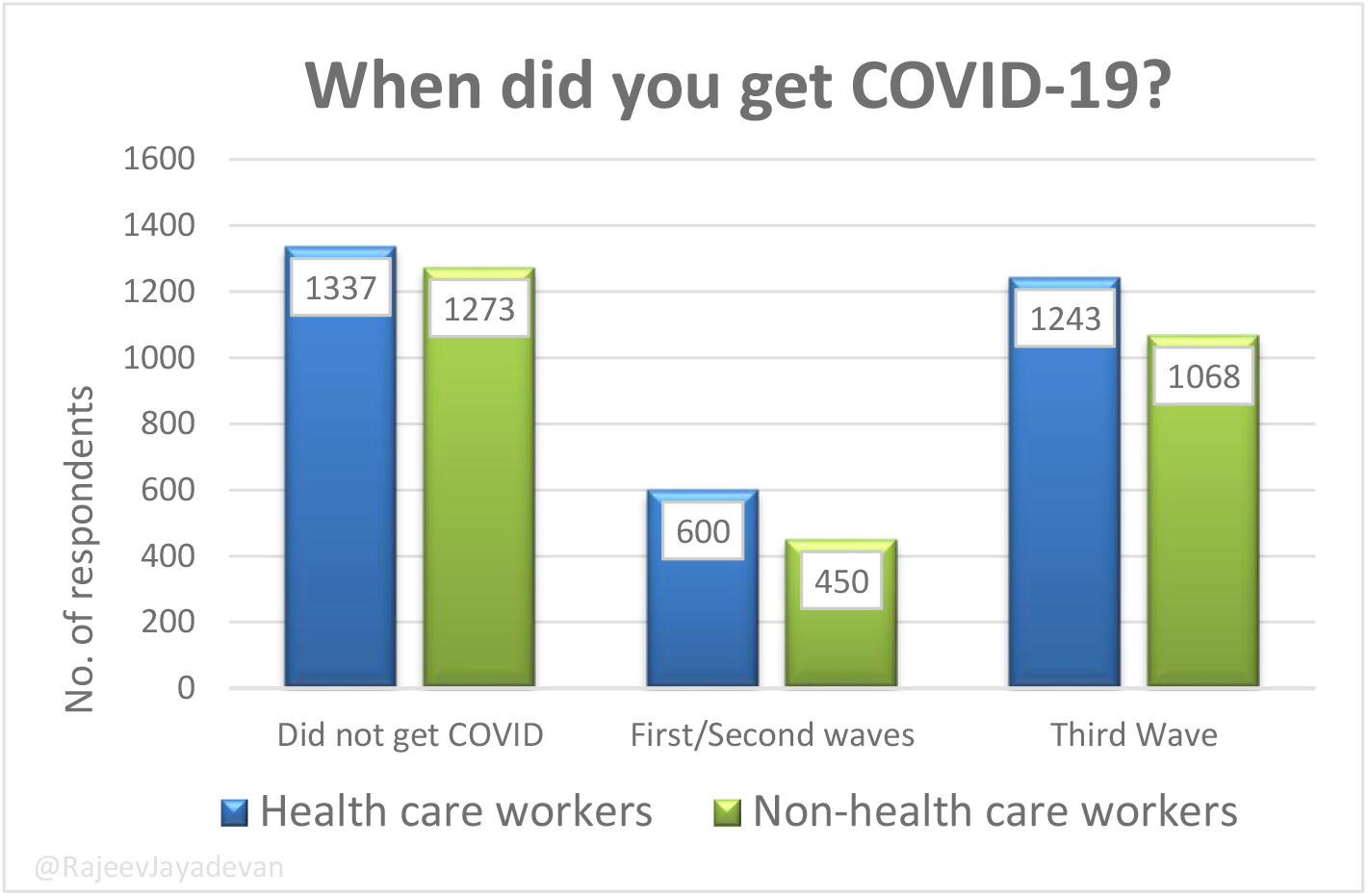
Timing of COVID-19.

**Figure 2:**
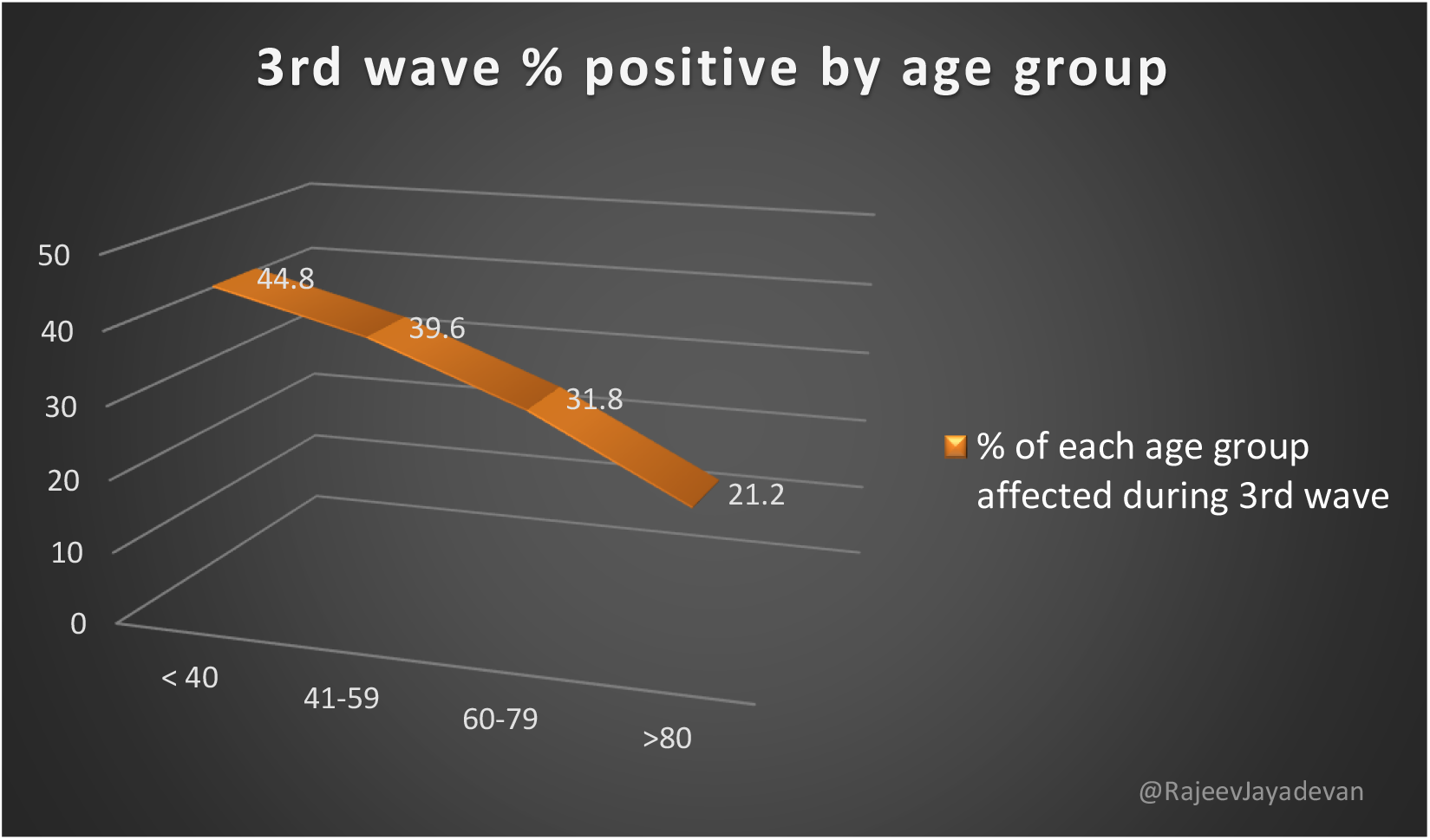
The percentage of people in each age group who were affected by the 3^rd^ wave

### 3. Severity of the third wave

Among the 2311 who were infected during the third wave, 4.8% were asymptomatic while 53% had mild symptoms. Moderate severity was reported by 41.5%, while 0.69% had severe COVID-19.

**Figure 3:**
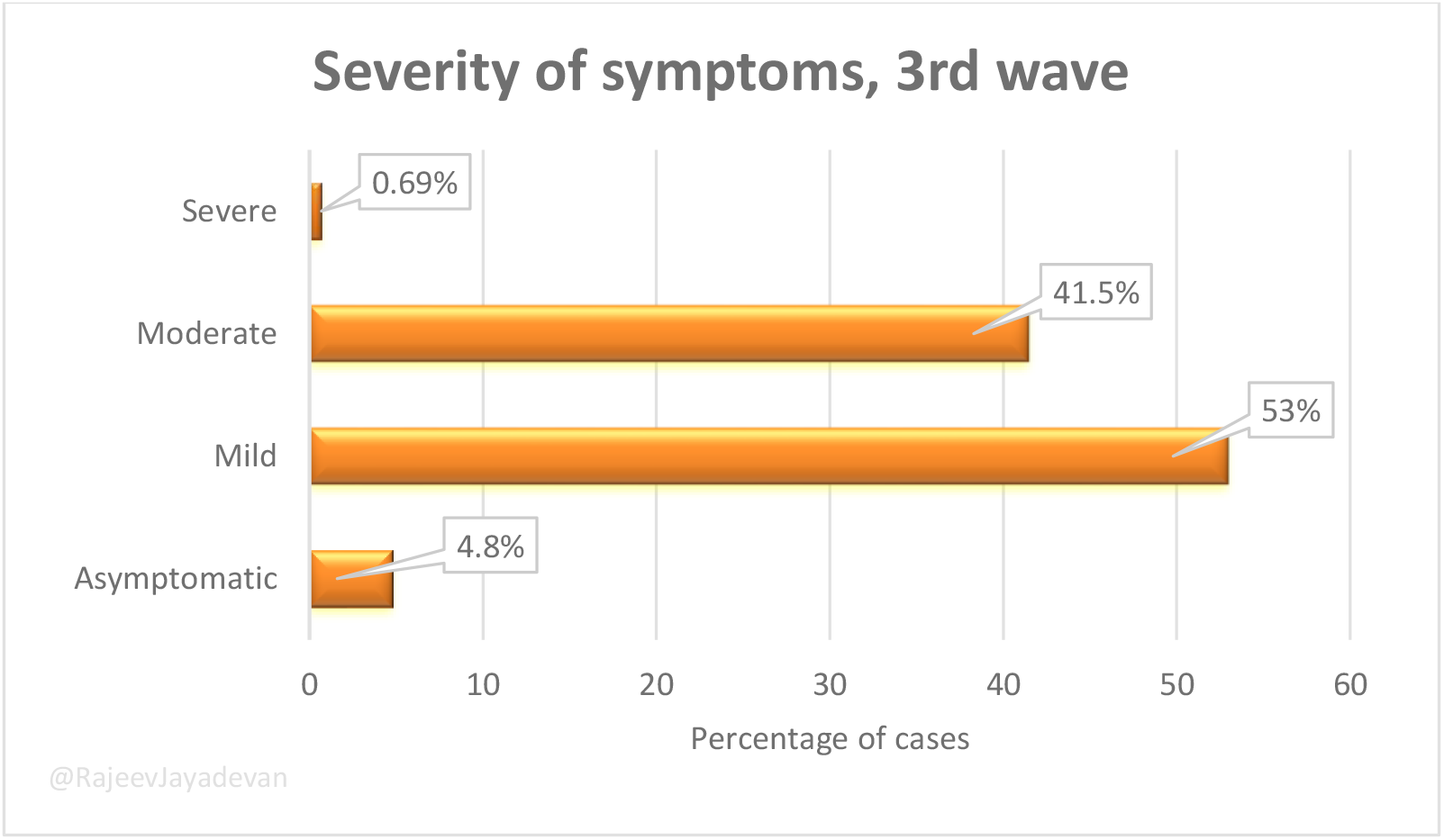
Symptom severity profile of 3^rd^ wave in India

### 4. Number of times infected, COVID-19

2868/3361 (85%) were infected only once. 454 (14%) had it twice. 26 people (0.8%) reported being infected 3 times. Two people reported being infected four times, while 12 (0.4%) said they had COVID-19 five times.

**Figure 4:**
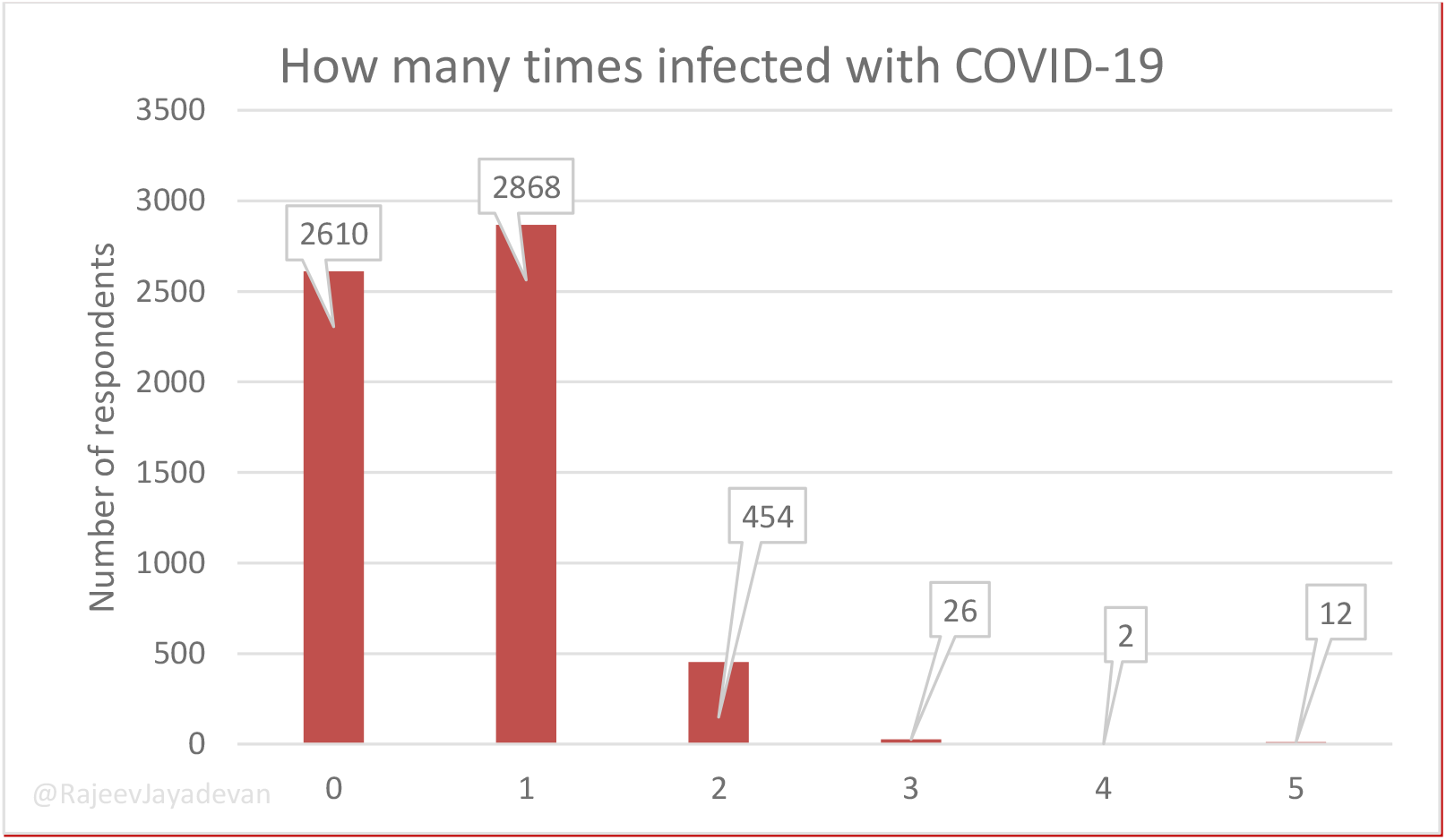
Number of times a person was infected

### 5. Profile of vaccine use

86.4% (5157) used Covishield, 8.8% (523) took Covaxin, 0.3% (20) Sputnik V and 3.1% (188) took others. 1.4% (83) were unvaccinated.

### 6. 3^rd^ wave infection pattern across vaccines, used as primary series

#### 1. Covishield

Among 5157 people who took it, 2010 (39%) reported COVID-19 during 3^rd^ wave

#### 2. Covaxin

Among 523 people who took it, 210 (40%) reported COVID-19 during 3^rd^ wave

**Figure 5:**
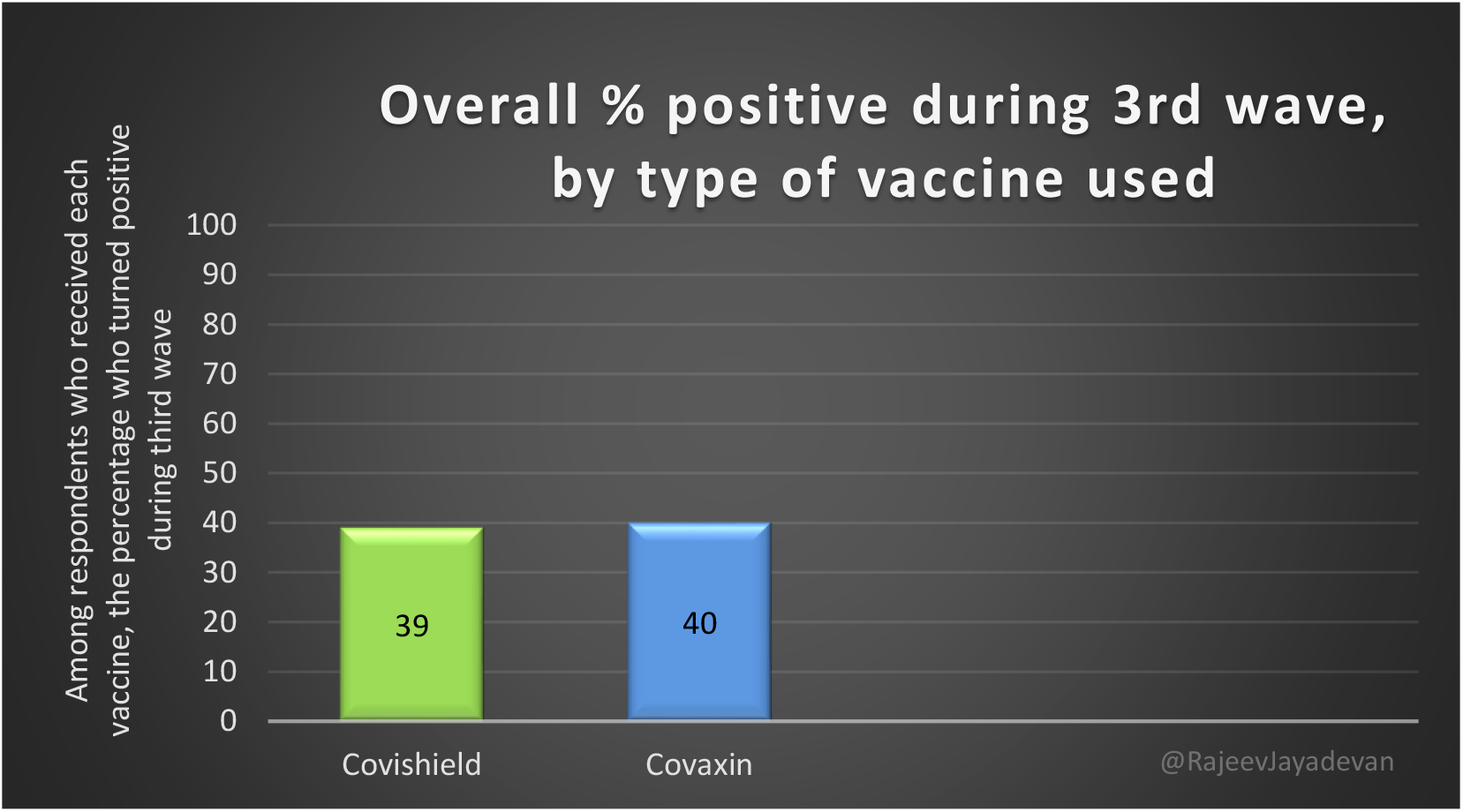
Percentage of vaccine primary series recipients who turned positive during the 3^rd^ wave

### 7. 3^rd^ wave infection pattern by vaccines, used as 3^rd^ dose

646 (30.2%) were infected out of 2140 who took Covishield 3^rd^ dose

38 (30.1%) were infected out of 126 who took Covaxin 3^rd^ dose

32 (29%) were infected out of 109 among other types of vaccine

**Figure 6:**
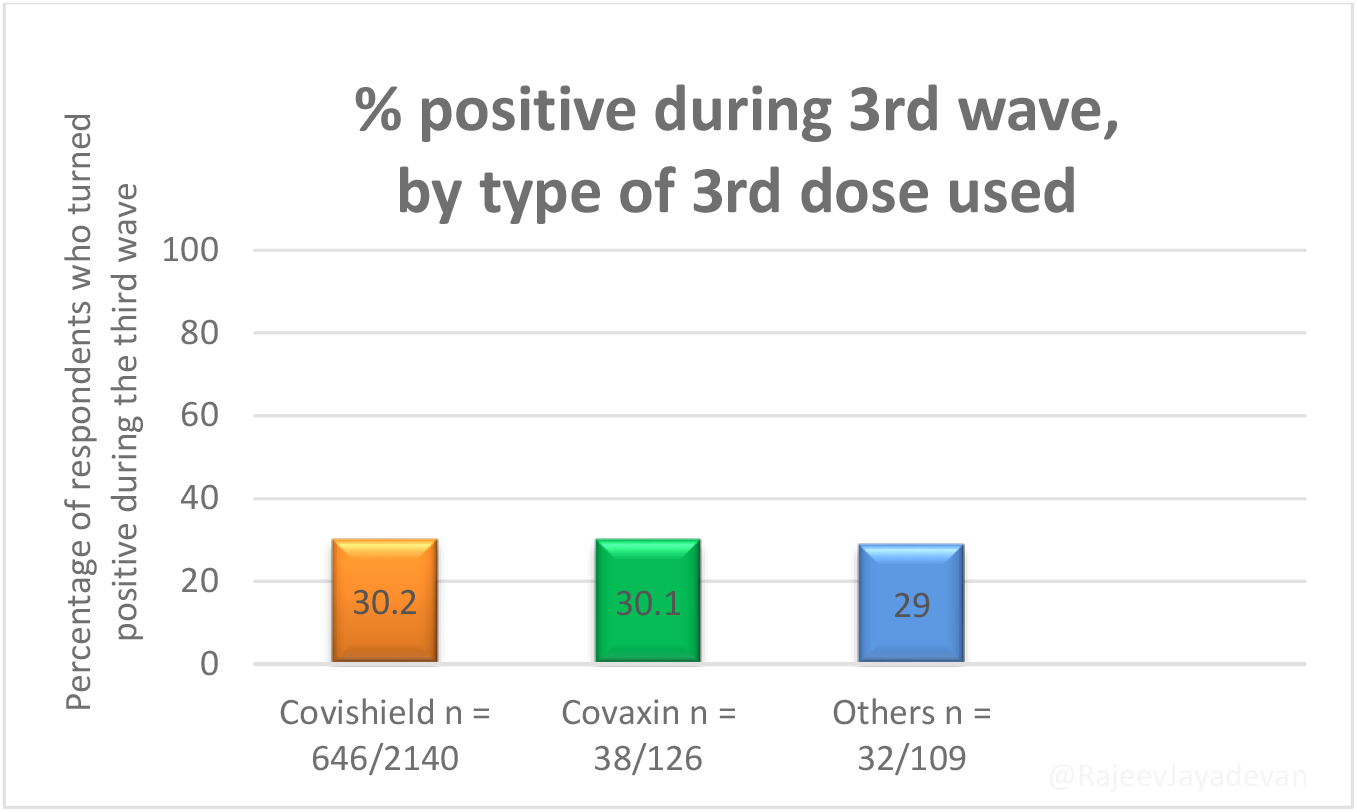
Percentage of 3rd dose vaccine recipients who turned positive in 3^rd^ wave

### 8. 3^rd^ dose uptake among respondents

2383 out of 5971 (40%) respondents took 3^rd^ dose

### 9. 3^rd^ dose uptake among healthcare workers

1701/3180 (53.5%) took 3^rd^ dose

Healthcare workers were more likely to receive 3^rd^ dose. This was expected because of the prioritisation during 3^rd^ rollout.

### 10. 3^rd^ dose uptake among non-healthcare workers

682/2791 (24.4%) took 3^rd^ dose

### 11. 3^rd^ wave positive rates among those who got 3^rd^ dose, versus those who did not take 3^rd^ dose

Of the 2383 people who took 3^rd^ dose, 716 (30%) reported positive during 3^rd^ wave

Among 3505 vaccinated people who did not take it, 1577 (45%) reported positive during 3^rd^ wave

**Figure 7A:**
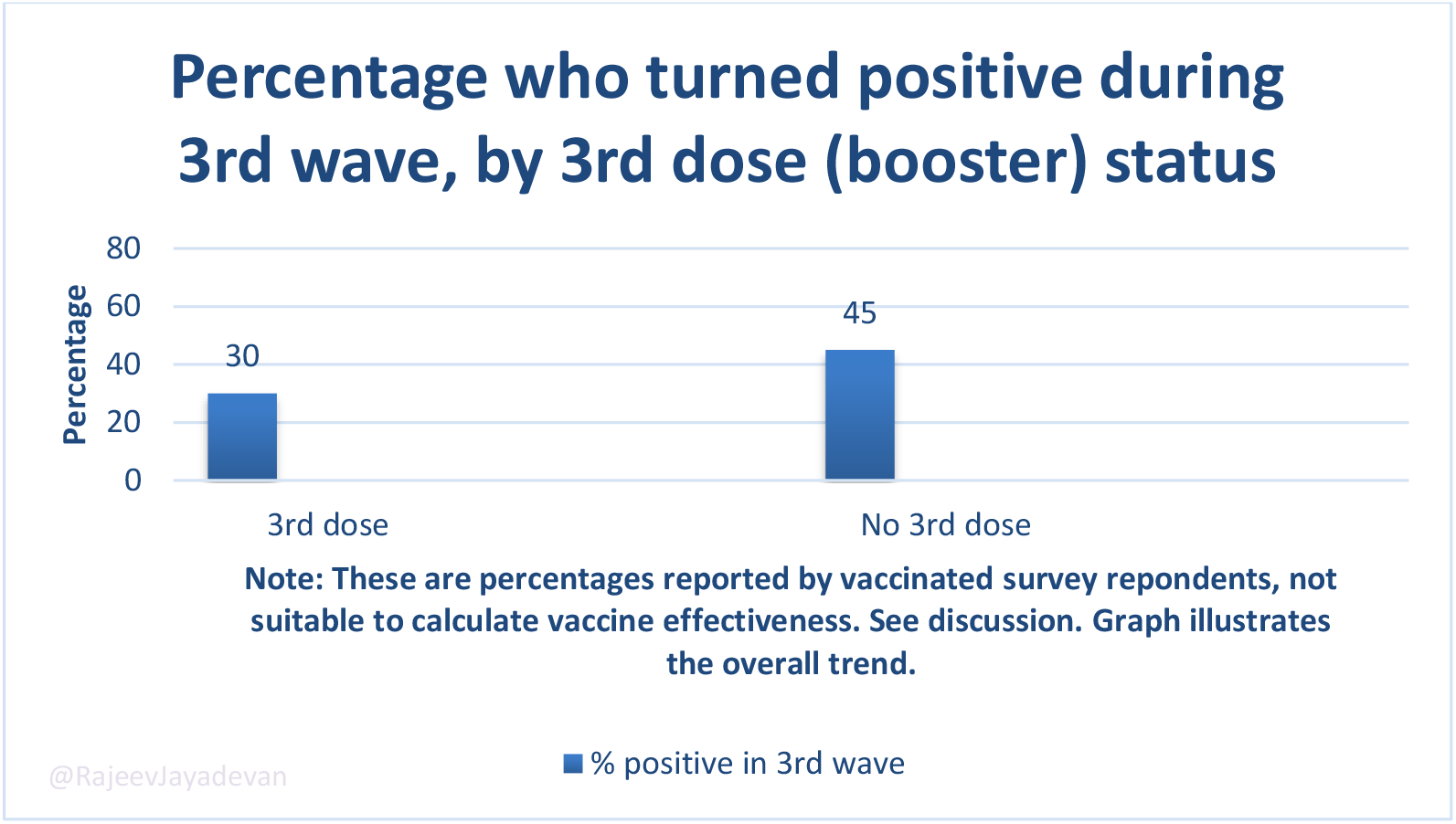
Third wave positivity according to whether 3^rd^ dose was taken or not

**Figure 7B:**
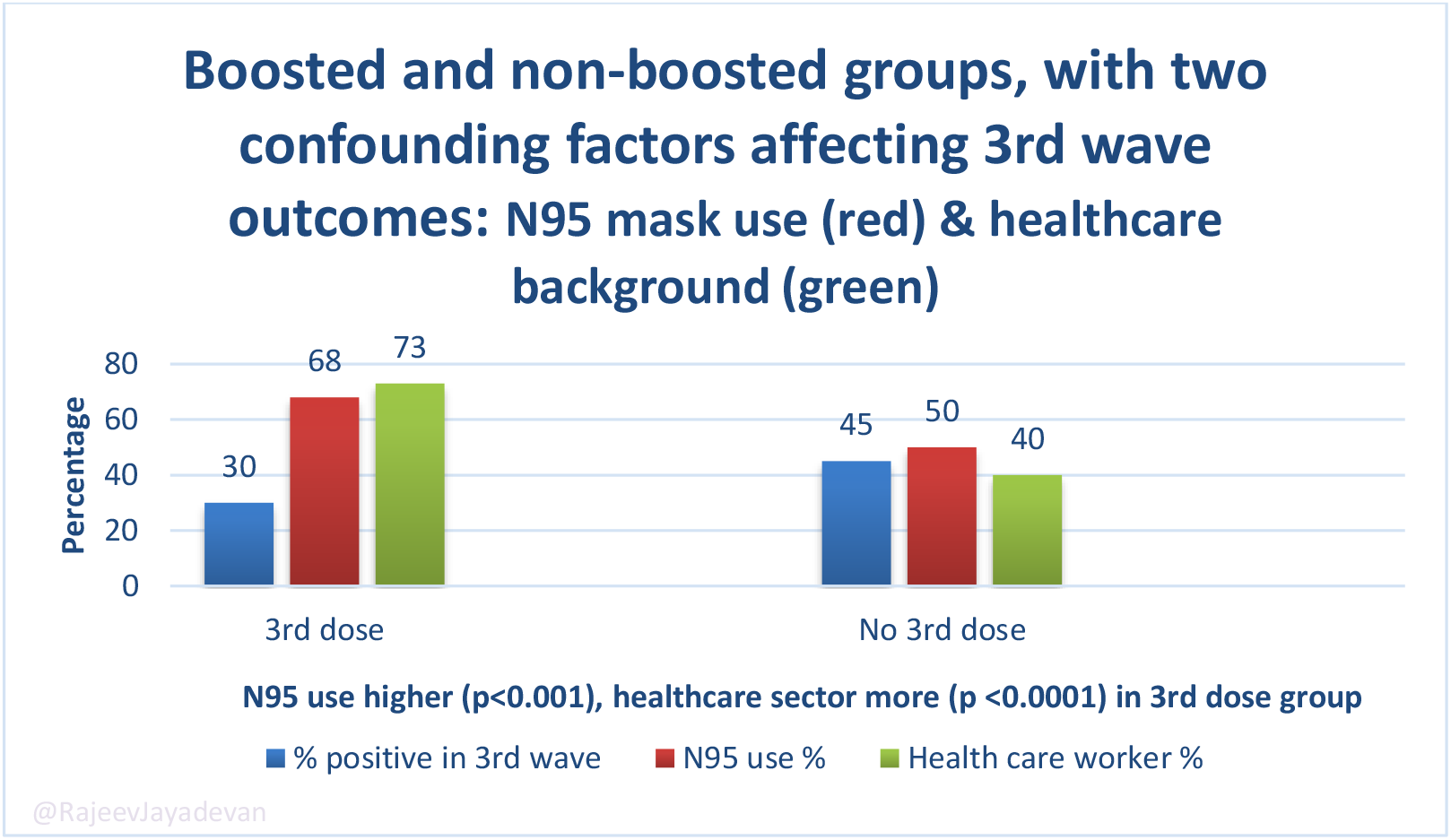
Comparison of confounding factors that could have affected the outcomes of 3^rd^ dose

### 12. 3^rd^ wave COVID-19 incidence, based on the time gap since 2^nd^ dose

#### 1. Analysis based on gap since 2^nd^ dose alone (some have taken 3^rd^ dose)

Among the recently vaccinated group (<1 month ago), only 27% (59/221) were positive in the 3^rd^ wave

Among the intermediate group, 37.3% (474/1270) were positive in the 3^rd^ wave

Among the > 6 month-gap group, 40% (1719/4267) were positive in the 3^rd^ wave

#### 2. Analysis after excluding those who also took the 3^rd^ dose from the above group

Among the recently vaccinated group (<1 month ago), only 27% (23/86) were positive in the 3^rd^ wave

Among the intermediate group, 38% (377/995) were positive in the 3^rd^ wave

Among the > 6 month-gap group, 49% (1147/2350) were positive in the 3^rd^ wave

This indicates that infection was more common among those who had a longer gap after the 2^nd^ dose. In other words, protection from infection appeared to be linked with *how recent* the last vaccine dose was.

**Figure 8 A:**
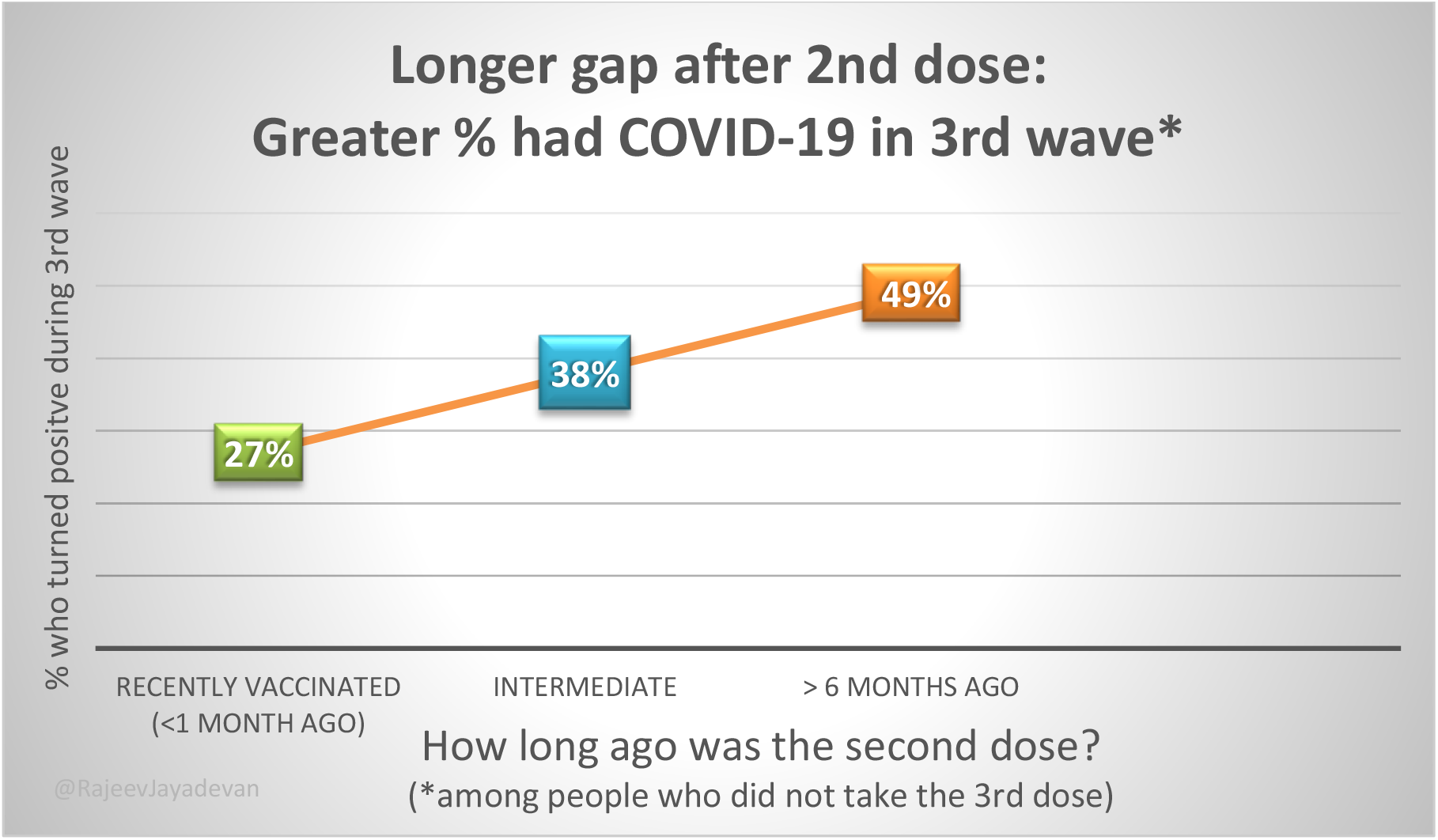
3rd wave positivity according to gap since receiving 2^nd^ dose, among those who did not take 3^rd^ dose

### 13. 3^rd^ dose outcome, based on gap since 2^nd^ dose

We looked at people who took the third dose after varying gaps from the second dose. They were categorized into three groups:

1. Recent: those who took 3^rd^ dose within 1 month of their second dose,
2. Intermediate: 3^rd^ dose taken after 2-5 months gap
3. Late: 3^rd^ dose taken >6 months after 2^nd^ dose

In these three groups, we assessed the impact of the 3^rd^ dose, as measured by the difference in 3^rd^ wave positivity rates. We found that a 3^rd^ dose given early or during the intermediate period made no difference to the baseline infection rate. But when the gap was > 6 months after the second dose, taking a 3^rd^ dose generated a 19% lower incidence of COVID-19 in the 3^rd^ wave.

**Figure 8 B:**
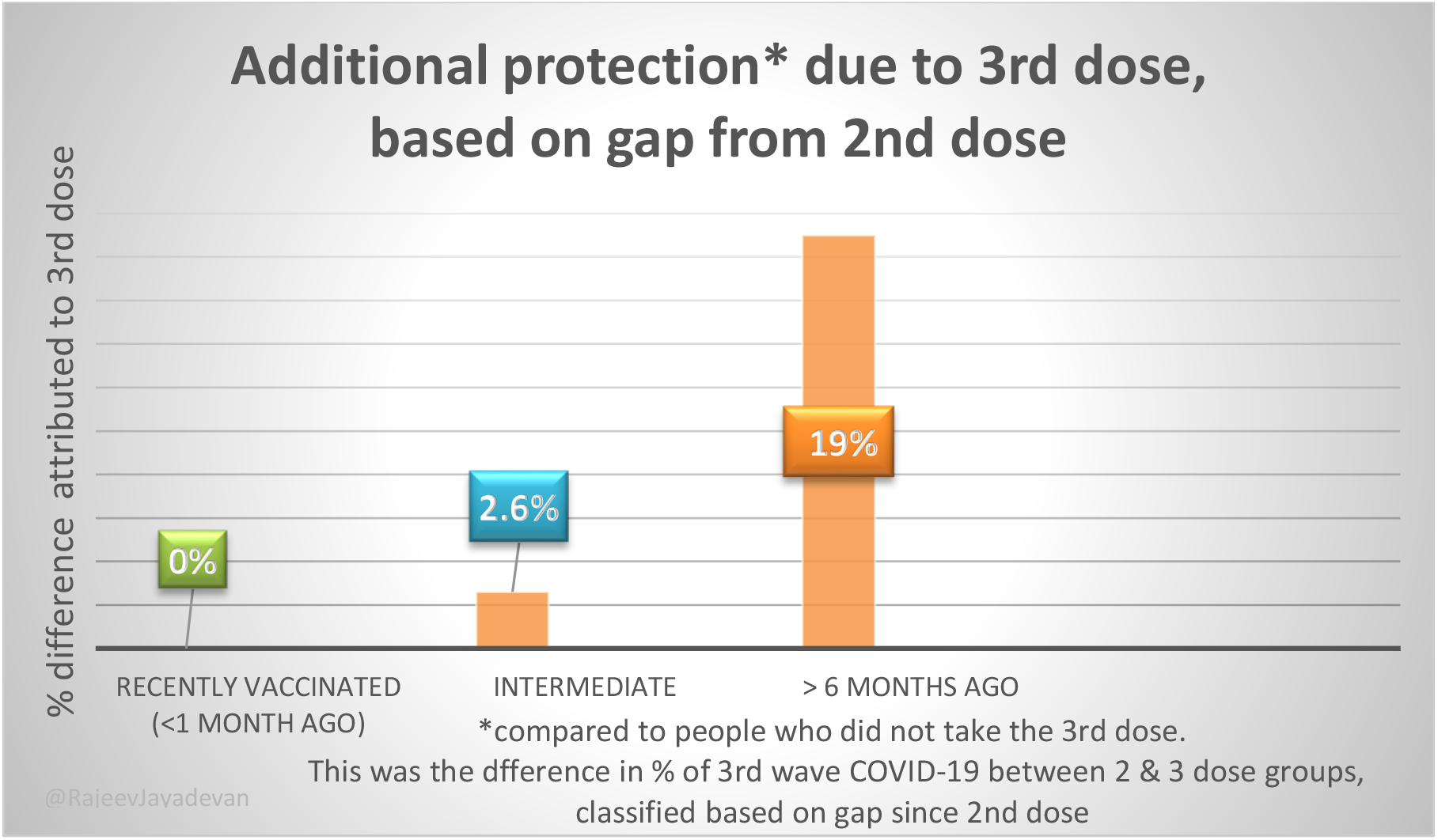
Difference in percentage of infection rates between 3 and 2 dose groups, classified on the basis of gap between 2^nd^ and 3^rd^ doses. The longer the gap, the greater the additional protection observed.

### 14. 3^rd^ wave infections after sufficient time elapsed since 3^rd^ dose

Of the 716, only 667 provided complete information on timing

Among 667 cases who had 3^rd^ dose breakthrough infection (that is, COVID-19 occurring in spite of taking a 3^rd^ dose), 516 (77%) got infected at least 2 weeks after the third dose.

### 15. Severity of 3^rd^ wave breakthrough infections

Among those who had a 3^rd^ dose, 716 got infected during 3^rd^ wave

- Asymptomatic 3%
- Mild 58.5%
- Moderate 37%
- Severe 0.3%

Among those who were vaccinated, but did not receive 3^rd^ dose, 1577 got infected during 3^rd^ wave

- Asymptomatic 5%
- Mild 50.8%
- Moderate 43.4%
- Severe 0.76%

**Figure 9:**
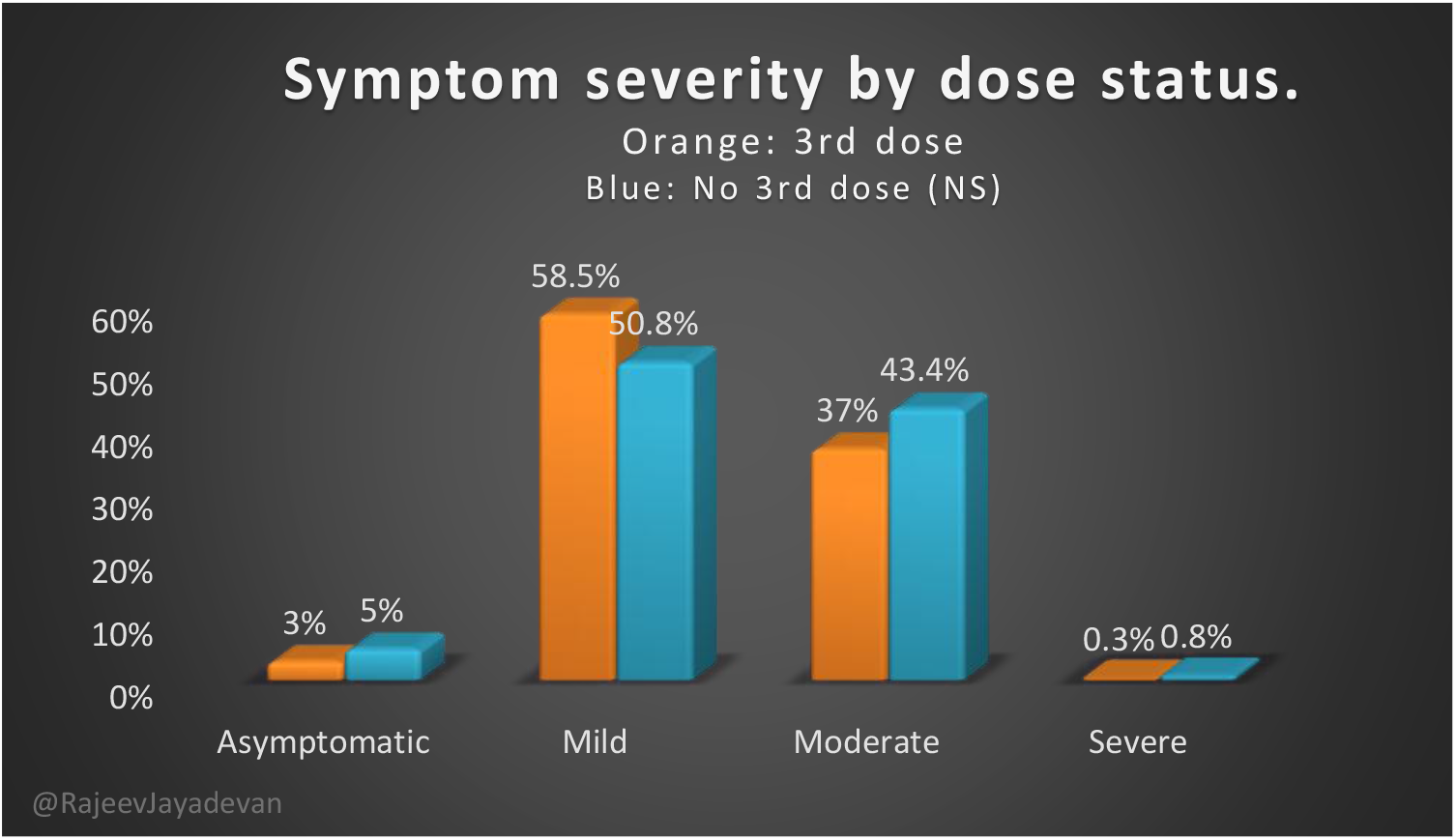
3^rd^ wave symptom severity profile classified according to whether 3^rd^ dose was taken or not

The severity profile of 3rd wave symptoms among vaccinated people who took 3rd dose and those who did not, were similar. The minor differences shown in the graph were not significant.

**Figure 10:**
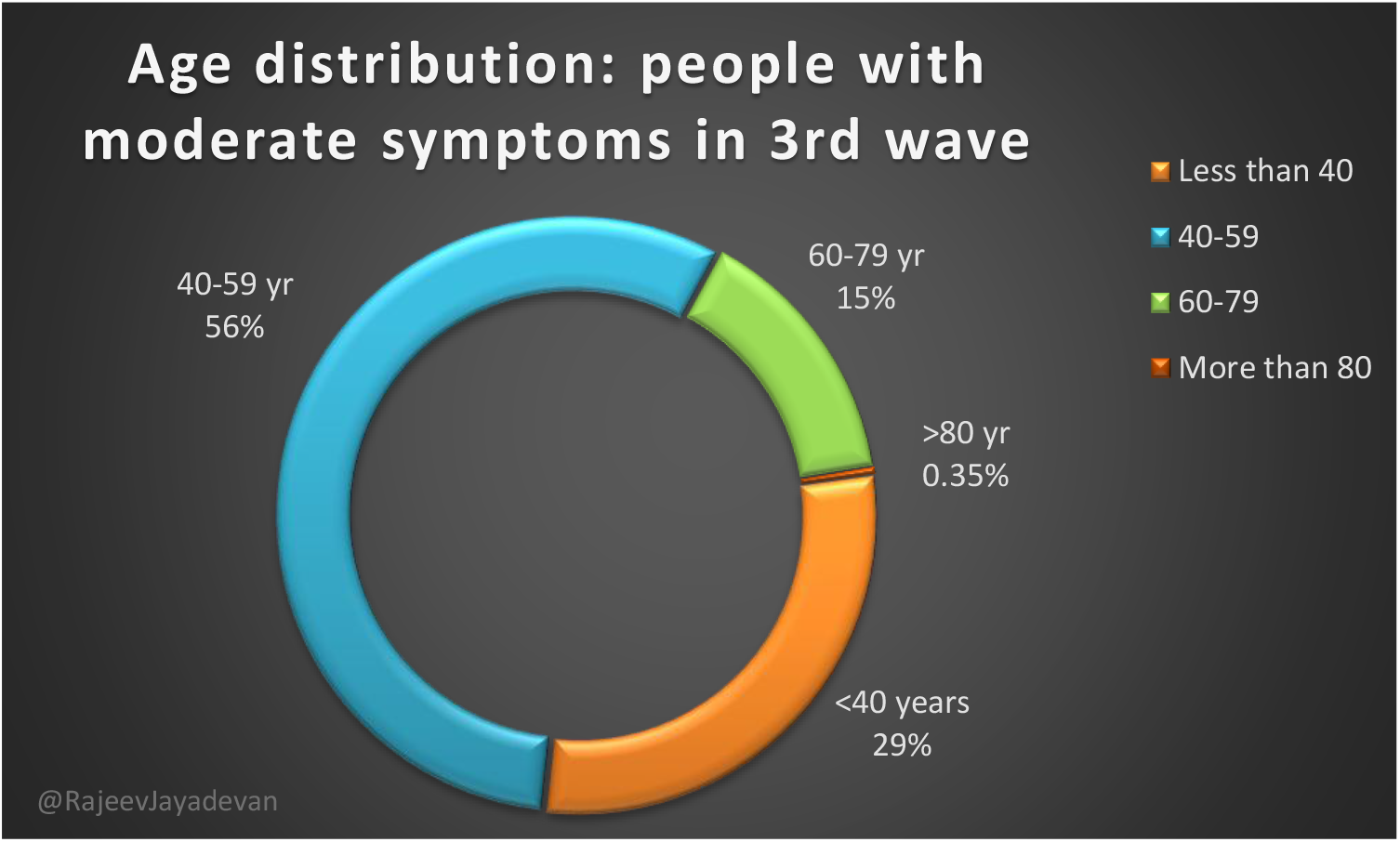
Age of those who suffered moderate severity of COVID-19 during 3^rd^ wave, expressed as % of total respondents. Although the chance of testing positive was higher among <40, the total number of respondents in the 40-59 age segment was greater.

### 16. Public opinion: “Do you think a precautionary dose is helpful?”

Overall, 2104/5971 (35.2%) believed it is helpful

3347 (56%) were unsure

520 (8.7%) did not think it was helpful

**Figure 11:**
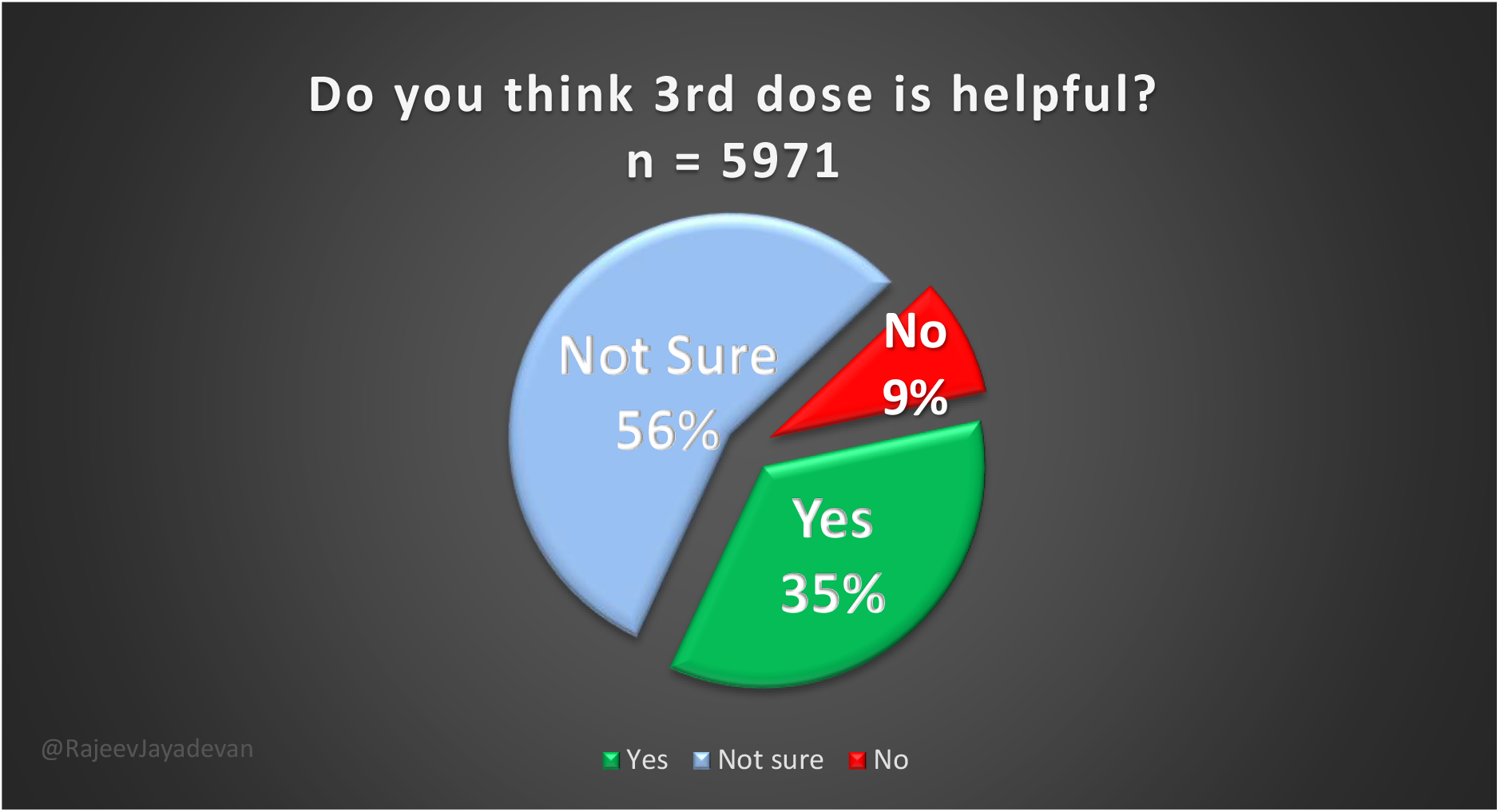
Public opinion about precautionary dose

### 17. Opinion among those 2311 infected in 3^rd^ wave

29% said helpful (671/2311)

58% were not sure (1345/2311)

12.7% said not helpful (295/2311)

### 18. Opinion among those 3660 who were not infected in 3^rd^ wave

39% said helpful (1433/3660)

55% were not sure (2002/3660)

6.1% said not helpful (225/3660)

**Figure 12:**
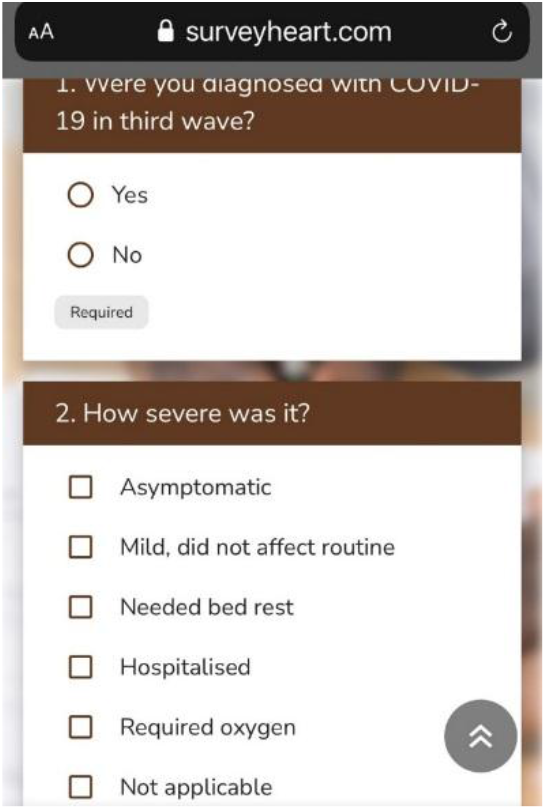
Sample of questions sent to assess severity of COVID-19 among respondents. For question no.2, more than one of the six choices could be selected, depending on individual severity. They were later classified as asymptomatic, mild, moderate and severe.

- Asymptomatic: option 1
- Mild: option 2
- Moderate: options 3 or 4, but not 5
- Severe: must include option 5

Those who were not infected could choose option 6.

**Figure 13:**
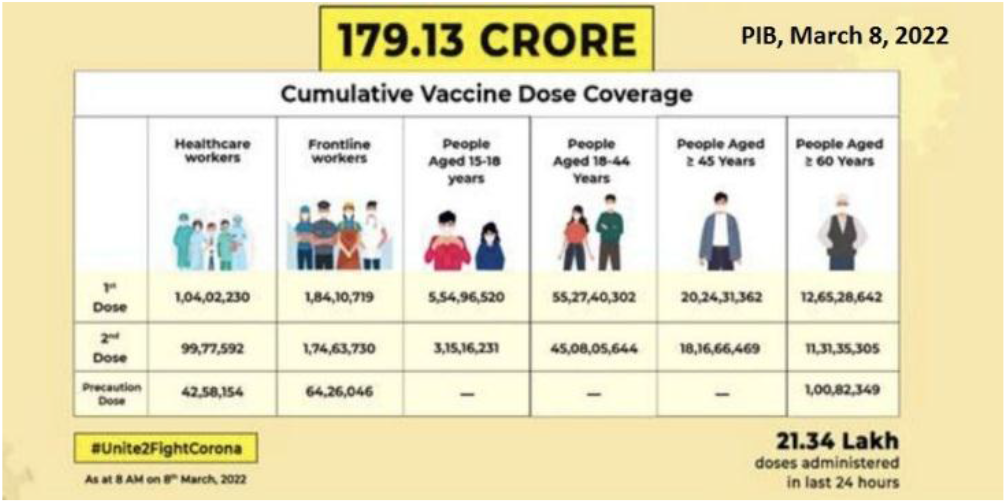
Vaccination status on March 8, 2022, by Press Information Bureau of India

## Discussion

The highlight of this cross-sectional survey was that 30% of those who took the third dose reported COVID-19 afterwards during the third wave. The high breakthrough infection rate following the 3^rd^ dose is remarkable because it occurred during the early period, where booster protection is expected to be maximum. This is consistent with reports elsewhere about Omicron being able to infect up to 64% of individuals who had recently received a 3^rd^ dose booster of mRNA vaccine (4).

We found that among the individuals who got 3^rd^ dose breakthrough infection, 77% got infected after sufficient time had passed after receiving the dose for antibody levels to be boosted (2 weeks or more). In other words, the high rate of infection observed was not because the 3^rd^ dose was taken too close to the third wave.

The survey indicates that despite a high baseline rate of vaccination among the respondents (98.6%), more people were affected in the 3^rd^ wave than during the initial two years, as reported elsewhere. (5) Among those infected during the 3^rd^ wave, symptoms of moderate severity were reported by 41.5%. This showcases not only the immune escape potential and transmissibility, but also the considerable morbidity and loss of productivity caused by Omicron.

For the purpose of classification, moderate disease included positive answers to the questions 1) needed bed rest and/or 2) hospitalisation with no oxygen required.

Less than 1% of those affected in the 3^rd^ wave reported severe disease, defined as those who required supplemental oxygen during hospitalisation (6). The percentage of severe disease during the 3^rd^ wave remained below 1% among all vaccinated individuals, whether they received the 3^rd^ dose or not.

The reported percentage of severe infections was only 0.6% in the 3^rd^ wave. It is noteworthy that 15% (3/20) of the severe cases were reinfections, and occurred among vaccinated individuals. This disproves the popular notion that reinfections are always milder than the first episode.

Younger adults <40 years of age had the highest positivity rate (Table 1, Figure 2). This was similar to the Omicron experience in South Africa (7) and could explain the relatively low severity observed.

**Table 1:** Demographic profile of the survey respondents

| Variables |  | COVID status, 3<sup>rd</sup> wave |  | Total | P value |
| --- | --- | --- | --- | --- | --- |
|  |  | Present | Absent |  |  |
| Age in years | Less than 40 | 637(44.8%) | 786(55.2%) | 2972(100%) | <0.0001* |
|  | 41-59 | 1178(39.6%) | 1794(60.4%) | 1423(100%) |  |
|  | 60-79 | 485(31.8%) | 1039(68.2%) | 1524(100%) |  |
|  | More than 80 | 11(21.2%) | 41(78.8%) | 52(100%) |  |
| Gender | Male | 1199(36.8%) | 2062(63.2%) | 3261(100%) | 0.002* |
|  | Female | 1110(41%) | 1597(59%) | 2707(100%) |  |
|  | Transgender | 2(66.7%) | 1(33.3%) | 3(100%) |  |

**Table 2:** 3^rd^ wave positivity by vaccine type, 3^rd^ dose, timing of 2nd dose, public opinion about precautionary dose

| Parameters |  | Covid Status |  | Total | P value |
| --- | --- | --- | --- | --- | --- |
|  |  | Present | Absent |  |  |
| Type of vaccine | Covishield | 2010(39%) | 3147(61%) | 5157 | 0.016* |
|  | Covaxin | 210(40.2%) | 313(59.8%) | 523 |  |
|  | Others | 67(35.6%) | 121(64.4%) | 188 |  |
| Timing of second dose | Less than 1 month ago | 23(26.7%) | 63(73.3%) | 86 | <0.0001* |
|  | 2-5 months | 377(37.9%) | 618(62.1%) | 995 |  |
|  | More than 6 months | 1147(48.8%) | 1203(51.2%) | 2350 |  |
| Status of precautionary dose | Yes | 716(30%) | 1667(70%) | 2383 | <0.0001* |
|  | No | 1595 | 1993 | 3588 |  |
| Is precautionary dose helpful? | Yes | 671 | 1433 | 2104 | <0.0001* |
|  | No | 295 | 225 | 520 |  |
|  | Not sure | 1345 | 2002 | 3347 |  |

Given equal baseline vaccination coverage among the respondents, reasons for this age gradient during the 3^rd^ wave could be greater mobility and social mingling among younger adults, and additional precautionary measures taken by older individuals.

The reported 3^rd^ wave infection rate among those who did not take the 3^rd^ dose was higher at 44.5%. The trend suggests that infection rate was lower among those who took the third dose (Figure 7a). At the same time, those who did not take the 3^rd^ dose had a similar overall severity profile to those who took the 3^rd^ dose. (Figure 9)

Although it is conceivable that the reduced infection rate was due to the effect of the 3^rd^ dose, it is noteworthy that the 3^rd^ dose uptake among healthcare workers (53.5%) was more than double that of those of non-healthcare background (24.4%). Among healthcare workers, factors such as superior mask use, knowledge and adherence to airborne infection preventive measures, and the downstream effects of prior exposure could have also reduced the rate of infection. For instance, healthcare workers in our survey were more likely to wear an N95 mask (78%) compared to non-healthcare (46%).

Accordingly, we found that the 3^rd^ dose group had significantly higher use of N95 masks (68%) than those who did not take 3^rd^ dose (50%), p <0.001. The 3^rd^ dose group also had a higher percentage of healthcare workers (73%) compared to the non-boosted group (40%), p <0.0001. (Figure 7b)

Testing rates could be lower among people who took the 3^rd^ dose due to greater confidence levels. Thus, asymptomatic or mildly symptomatic cases could have been missed.

In other words, those who received the 3^rd^ dose had other protective factors too working in their favour.

Unlike a randomised trial or test-negative case-control study where such confounders can be controlled for, a survey is not a suitable tool to directly compare incidence rates, particularly between dissimilar groups of respondents.

In response to the question about whether they felt the precautionary dose was helpful, only 35% felt it was helpful, 9% believed it was not helpful, while 56% were unsure. That two-thirds of the respondents were either unsure or disapproving of the 3^rd^ dose is a significant observation, considering this group has a 98.6% vaccine acceptance rate prior.

Those who were infected during the third wave despite the 3^rd^ dose had lower confidence in the 3^rd^ dose, only 29% believed it was helpful. The continued trust was because some of them believed the 3^rd^ dose made their disease milder.

The following were the main reasons people gave for taking the 3rd dose:

1) ‘vaccine boosts immunity’ 2) ‘vaccine made disease milder’ 3) ‘vaccine had previously helped prevent infection in spite of known exposure’.

The chief reason that people mentioned for not taking the 3rd dose was that infections were being commonly reported after the third dose. Several respondents also shared their personal experience of getting infected despite the 3^rd^ dose.

Other stated reasons for not taking 3^rd^ dose were: 1) belief that prior infection would be protective 2) ‘lack of enough evidence’ 5) adverse experience with prior doses of vaccine 6) concern that mutations have altered the virus since the vaccine was originally made 7) ‘two doses were enough’ and 8) ‘waiting for mix-and-match vaccines’

Although the survey did not specifically include questions about adverse reactions, 3 of the 795 comments mentioned minor side effects following 3^rd^ dose. They were transient tiredness, fever, body ache and throat discomfort. We had previously reported the adverse effect profile of the primary vaccination series in India (8).

We found that a sizable proportion of people (44%) reported no known history of COVID-19 so far. It is possible that many in this group had asymptomatic infections, or had not been tested. Since the respondents belonged to diverse demographic groups, we believe that we got a true representation of the community from a wide geographic distribution.

Most people did not know the exact source of picking up the infection, but workplace (medical outpatient clinic for healthcare workers), spouse and child were the most frequently cited sources.

A large number of people (15% of those with a history) reported having COVID-19 more than once. Among them, 454 people had it twice, 26 people thrice and a few individuals reported up to 5 times. This is consistent with observations elsewhere about reinfections being common (9). The reinfection percentage of 15% is likely to be an underestimate because several respondents clarified that in subsequent episodes with compatible symptoms, testing was not always done. Unfortunately, it is not possible to verify each one of these cases because the survey is entirely self-reported. It is likely that a few of these episodes were self-diagnosed.

The 3^rd^ wave infection rates were almost identical (39 and 40%) across the two main vaccines used in India, which are Covishield (adenovirus vector) and Covaxin (inactivated), when used as primary series. When these vaccines were used as 3^rd^ dose, the third wave infection rates were identical at 30%. Among those few respondents who had taken other vaccines (breakdown not available due to survey constraints), the third wave infection rate was 29%.

Using the survey, we tried to determine whether the infection rate depended on how recent the vaccination was. We found that the longer the gap after the second dose, the greater the likelihood of becoming infected during the 3^rd^ wave. Accordingly, the percentage of 3^rd^ wave infection was only 27% among people who had recently received a second dose (Figure 8). This was nearly the same as 30% among those who recently received a 3^rd^ dose. This suggests that proximity to the most recent vaccine dose - whether it was the second or the third - is an important determinant of protection from infection. This is consistent with reports that for corona viruses, protection from reinfection is short-lived (10).

We also found that among those who had taken their second dose, adding a third dose without sufficient gap did not confer any additional protection from infection (Figure 8b). When the 3^rd^ dose was give after a 6 month gap, there was a 19% reduction in the risk of infection. This suggests that frequent vaccine doses will not add anything further to pre-existing protection from infection.

Among those who reported severe disease, 15% were reinfections. We contacted individuals who had repeated episodes of COVID-19, and asked them whether the second episode was milder. From the limited number of individuals that could be reached, no consistent pattern was apparent; some said the second bout was more severe, a few said that they were equally severe requiring several days of rest, others said it was milder. This is an area that needs further study, especially considering the seemingly endless risk of reinfections in the years to come. If large studies of reinfections show that subsequent bouts cause more severe disease, mitigation measures will need to be upgraded.

The strength of the study was the large number of respondents and the diverse demography, which are reassuring of a balanced sample. That 44% of respondents did not report COVID-19 was an indication that the results were not skewed in any direction, and presented a broad picture of the general population as the country went through its third wave. The comments section served two purposes. It encouraged anonymous sharing of personal views without constraint, thus generating a positive user experience, indirectly leading to greater survey referrals. The descriptive comments entered by 795 respondents also helped corroborate the main survey findings.

The limitations of our study are that because it was a survey, all the data provided could not be independently verified. All positive and negative cases were based on self-reported information. Besides, limitations in language proficiency in individual cases could have affected the quality of the response. While analysing the data, we did however talk with several respondents by phone, where contact information was provided. A significant limitation was that the number of unvaccinated individuals was small, constituting only 1.4% of the total number of respondents. Therefore, a meaningful comparison of their disease profile was not possible.

A cross-sectional survey is a descriptive research tool that provides a broad snapshot of society at a given time-point, and is not to be equated with a case-control study or a randomised trial that are able to select the participants and oversee data collection. Although useful to study disease patterns across large sections of the population, a survey is therefore not a method that can be used to calculate effectiveness of interventions like booster doses.

## Conclusions

Among those who took the third dose, nearly one-third reported COVID-19 during the third wave. Although less than one percent had severe symptoms during the 3^rd^ wave, 41.5% reported symptoms of moderate severity. Younger adults were preferentially affected. There was no difference between vaccine type and 3^rd^ wave infection rate.

Among vaccinated individuals who had not received 3^rd^ dose, 45% reported COVID-19 during the 3^rd^ wave. However, this group had significantly lower use of N95 masks (50%) than the 3^rd^ dose group (68%), which may have contributed to higher infection rates.

The 3^rd^ wave symptom severity profile was similar among those who took 3^rd^ dose and those who had primary vaccination series.

Repeated bouts of infection were reported by 15%, this number is likely to be an underestimate due to diminishing testing rates. Reinfections were not necessarily milder.

A longer gap after the second dose correlated with a greater chance of being infected in the 3^rd^ wave. Among those who had recently received their second dose, only 27% (59/221) were positive in the 3^rd^ wave, which was about the same as that following the 3^rd^ dose (30%). This suggests that infection was less likely among those who had recently received a vaccine dose.

The 3^rd^ dose, given too close to the second dose, made no difference in the infection rate.

Although the respondents were 98.6% vaccinated at baseline, there was considerable uncertainty (65%) amongst them about the benefit of a 3^rd^ dose.

The high breakthrough infection rate implies that vaccination as a standalone strategy will not be enough in the days to come. Multiple strategies of protection will likely be required long-term from a public health perspective.

## Data Availability

All data produced in the present work are contained in the manuscript

